# Screening of plasma IL-6 and IL-17 in Bangladeshi lung cancer patients

**DOI:** 10.1101/2022.03.27.22272998

**Authors:** Manik Chandra Shill, Bisshojit Biswas, Moriam Islam, Sharmin Sultana Rima, Farhana Afrin Ferdausi, Qamruzzaman Chowdhury, Hasan Mahmud Reza, Asim Kumar Bepari

## Abstract

Cancer is the second leading cause of death globally, where most cancer deaths occur in low- and middle-income countries. Lung cancer is the most prevalent in men and the third most prevalent in women among all cancer types. Globally, 1.8 million new lung cancer cases were recorded in 2012, which increased to 2 million in 2018. Cancer disease burden can be diminished, and life expectancy profoundly increases when diagnosis and prognosis are made at an earlier stage. Although biopsy is the gold standard in cancer diagnosis, it is invasive, inconvenient, and expensive. Liquid biopsy, which identifies cancer biomarkers in blood, is an active area of cancer research. Recent evidence suggests that interleukins (ILs), a class of cytokines involved in diverse cellular processes such as inflammation, growth, and proliferation, play critical roles in cancer initiation, progression, and resistance to therapy. Interestingly, many studies found the association of plasma IL-6 and IL-17 levels with lung cancer prognosis. In this study, we analyzed plasma levels of IL-6 and IL-17 in lung cancer patients and healthy volunteers. We have also studied the demographic and medical records of the participants. Among the participants, 42.9% and 57.1% were female in the control group and the disease group, respectively. The age of the participants was 22-65 years, with a mean of 46.5 (SD, ± 19.7) years. Plasma IL-6 levels were strikingly different between healthy volunteers (mean ± SEM, 0.97 ± 0.15 pg/mL) and patients (mean ± SEM, 7.42 ± 1.45 pg/mL). Notably, the range was narrow in the control group (0.32-2.10 pg/mL) but wide in the disease group (0.42-23.20 pg/mL). Plasma IL-17 levels were slightly lower in the disease group (mean ± SEM, 9.40 ± 2.82 pg/mL) compared to the control group (mean ± SEM, 12.40 ± 4.41 pg/mL), although the difference was not statistically significant. Intriguingly, the mean IL-17 value reduced dramatically with chemotherapy, and further reduction occurred with radiotherapy in lung cancer patients. Together, our study supports the use of plasma IL-6 and IL-7 levels as prognostic markers in lung cancer.

## Background Information

Cancer is one of the biggest threats to public health worldwide, accounting for nearly 17% of global deaths. The scenario is notably worse in low- and middle-income countries where access to preventive, diagnostic, and treatment facilities is inadequate. In Bangladesh, approximately 10% of total deaths occur due to cancer. Although the rate of cancer deaths in Bangladeshi females shows a decreasing trend after 2005, the death rate is steady for males. According to a WHO report, the incidence of lung cancer is the highest among all cancer types in Bangladeshi males.

More than half of the cancer burden can be reduced by implementing preventive measures and early diagnosis. Tumor tissue biopsy is the gold standard in the diagnosis of cancers. However, in most cases, invasive diagnostic procedures are expensive and inconvenient. In this context, blood biomarkers or ‘liquid biopsy’ emerge as a potential tool for cancer diagnosis and prognosis.

Although inflammation is a physiological protective mechanism, inappropriate inflammatory responses have been implicated in many diseases. Inflammation has significant impacts on cancer initiation, growth, progression, and resistance to chemotherapy (Dmitrieva et al., 2016). Interleukins, a class of cytokines expressed in various cell types, including leukocytes, are critical regulators of inflammatory responses in carcinogenesis. Dmitrieva et al. (2016) reviewed the roles of interleukin-1 (IL-1) and IL-6 in inflammation and cancer (Dmitrieva et al., 2016). Tissue damage and oxidative stress activate the transcription factor NFκB in immune cells, inducing the generation of IL-1 and IL-6, which can promote oncogene expression. Levels of IL-1 alpha in cancer patients’ tumor tissues correlated with tumor regression, whereas IL-1 beta indicated an unfavorable prognosis. IL1 also generates other cytokines and proinflammatory mediations, including reactive oxygen species. Many studies found an association of IL-6 levels with tumor growth, metastasis, and survival in many cancer. Silva et al. (2017) observed a statistically significant inverse correlation of plasma IL-6 levels and overall survival of Bazilian non-small cell lung cancer (NSCLC) patients (Silva et al., 2017). Recent studies also suggest that IL-17 regulates lung cancer growth, progression, and metastasis by promoting angiogenesis and proliferation and downregulating apoptosis (Wu et al., 2016). In a meta-analysis involving 479 Chinese lung cancer patients, elevated IL-17 expression correlated with poor clinical outcomes (Wang et al., 2017).

In the present study, we evaluated plasma levels of IL-6 and IL-17 in Bangladeshi lung cancer patients.

## Materials and Methods

### Patients

In 2019, lung cancer patients were enrolled at a cancer hospital in Dhaka city, Bangladesh. Written informed consents were received from each participating patient before enrollment. Epidemiological data including age, weight, place of residence, smoking behavior, and the previous history of malignancy were collected from medical records and a short questionnaire. Clinical and histopathological data were obtained from available medical and pathological reports. Inclusion criteria for the patient group were the age of 18 years or more and confirmed diagnosis of lung cancer. Healthy individuals without any present or past cancer history were enrolled in the control group. This study was approved by the NSU Institutional Review Board/ Ethics Review Committee (IRB/ERC).

### Sample collection

In indisposable BD Vacutainer tubes containing 6% ethylenediaminetetraacetic acid (EDTA), five milliliters of peripheral blood). The whole blood will be centrifuged at 700 x g for 10 minutes. Blood plasma was then be further centrifuged at 2,000 x g for 10 minutes at 4 ºC. The plasma samples were aliquoted and stored at -80 °C until further use.

### Assay of plasma IL-6

The human IL-6 ELISA Kit (ab46027) was purchased from Abcam, and the assay was performed according to the manufacturer’s instructions. In brief, all reagents were prepared and equilibrated at room temperature before the experiment. We added 100 µl of each standard solution and samples to appropriate wells of the microplate. Next, 50 µl biotinylated anti-IL-6 antibody solution was added to each well. The plate was covered and incubated at room temperature for 1 hour. Subsequently, the wells were washed three times using the 1X wash buffer. We then applied Streptavidin-HRP solution into all wells and incubated it for 30 minutes at room temperature. After three washes, we added the Chromogen TMB substrate solution into each well. Incubation was continued for 20 minutes in the dark. Then, we terminated the color development by adding 100 µl stop reagent to each well. Absorbance was measured on a spectrophotometer at 450 nm.

### Assay of plasma IL-17

The human IL-17 ELISA Kit (ab119535) was obtained from Abcam. We performed the assay of plasma IL-17 following the manufacturer’s guidelines. All materials were equilibrated at room temperature before use. Working reagents were prepared as instructed. The microplate wells were washed twice with 400 µl wash buffer per well. Standard dilutions (100 µl)were pipetted into designated wells. Then, 50 µl sample diluent and 50 µl were added to sample wells. Biotin-conjugated anti-IL-17 antibody solution (50 µl) was added to all wells. The plate was incubated at room temperature for 2 hours. After the incubations, all wells were washed four times, and then 100 µl Streptavidin-HRP solution was pipetted into each well. The plate was further incubated for 1 hour at room temperature. All wells were then washed four times with the wash buffer, and 100 µl TMB substrate solution was added to each well. The color development was continued for 15 minutes in the dark at room temperature. The chromogenic reaction was stopped by adding 100 µl stop solution to each well. Absorbance was recorded on a spectrophotometer at 450 nm as the primary wavelength.

### Data Analysis

Data entry was done using an MS Excel worksheet. We also used MS Excel to analyze data and the generation of charts.

## Results and discussion

### Demographic data

We collected plasma samples and demographic and medicopathological data from 35 lung cancer patients (Disease group) and 19 healthy volunteers (Control group). Female participants were 15.8% and 11.4% in the control and disease groups, respectively (Figure 1).

**Figure 1:**
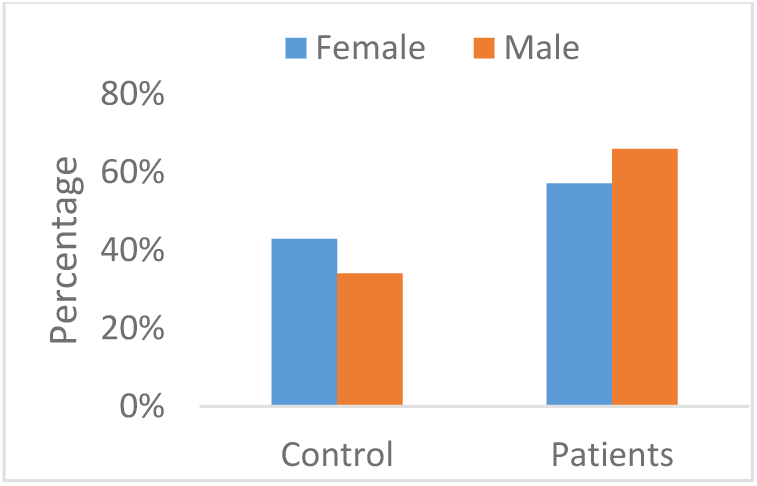
Gender distribution of participants.

The age range was 26-87 years in the patients and 22-32 years in healthy volunteers. Figure 2 shows the distribution of age groups of participants.

**Figure 2:**
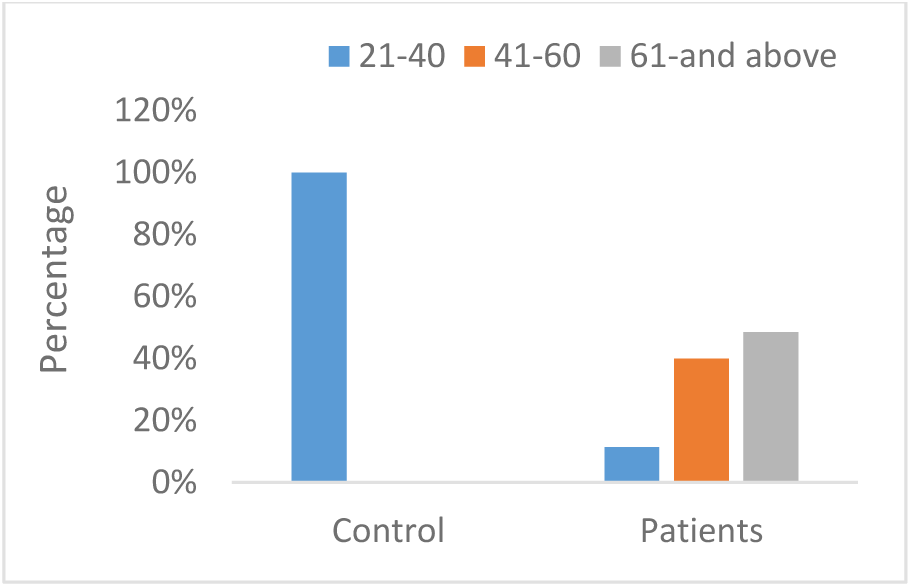
Age of participants.

**Figure 3:**
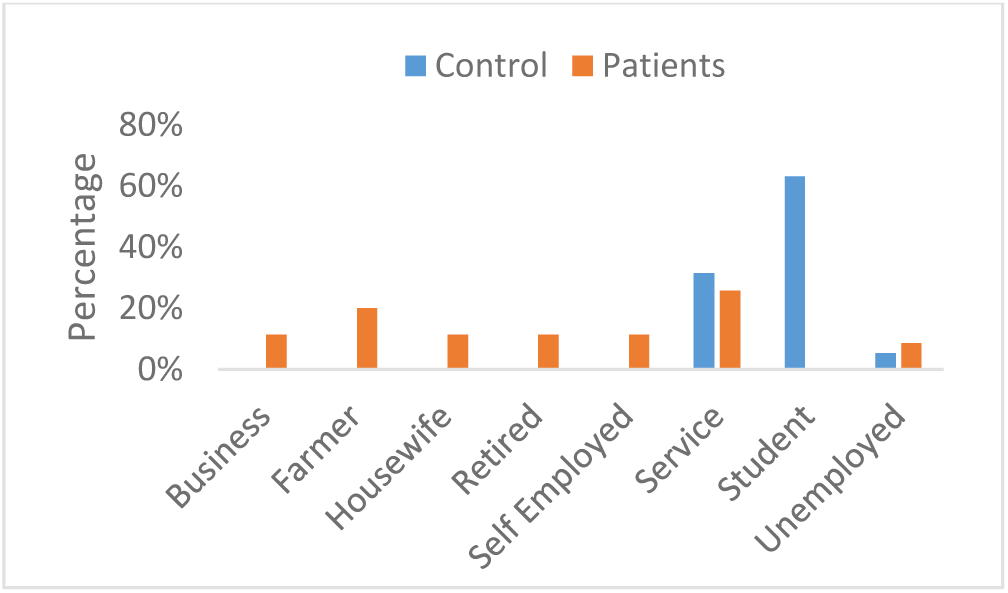
Profession of participants

The profession of lung cancer patients varied widely; however, the healthy volunteers were primarily students and service holders.

A family history of cancer (parents or siblings) was 21% in the control group and 25.7% in the disease group (Figure 4).

**Figure 4:**
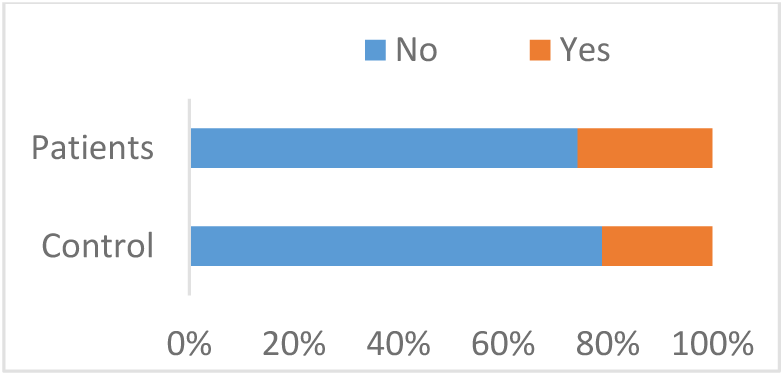
Family history of cancer.

We asked the participants about the level of stress they were experiencing. In both groups, the majority reported having a medium level of lifestyle-related stress (Figure 5).

**Figure 5:**
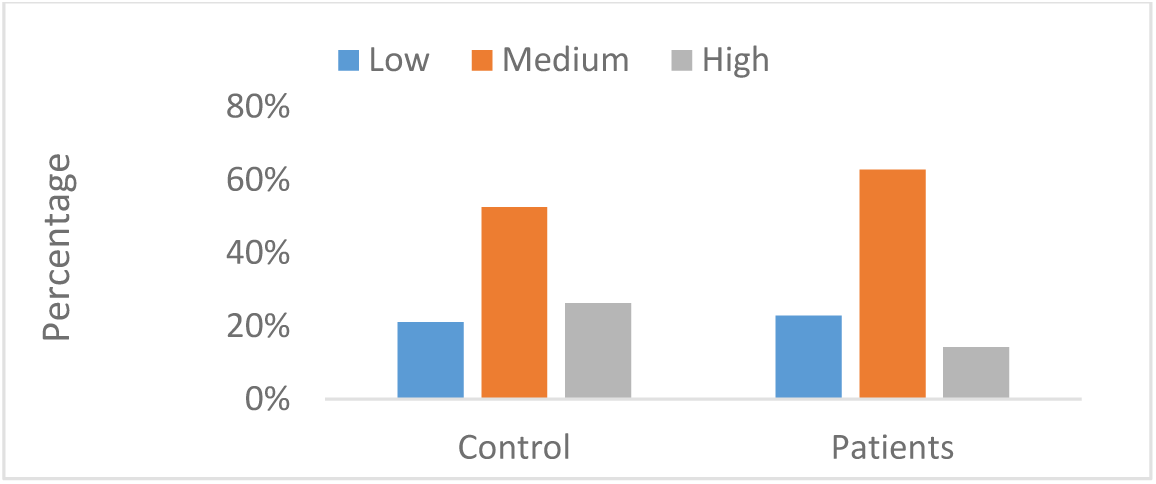
Stress levels in participants.

We collected information about the number of cigarettes smoked per day by the participants. We then categorized as non-smokers (no smoking), light-smokers (1-10 cigarettes/day), medium-smokers (11-30 cigarettes/day), and heavy-smokers (31 and more cigarettes/day) (Figure 6).

**Figure 6:**
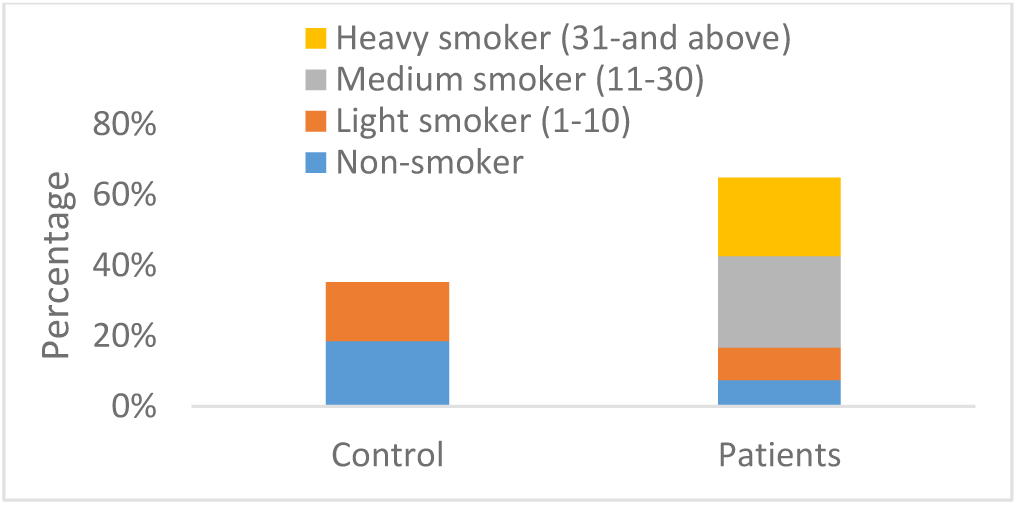
The smoking habit of participants.

Participants in the control group were primarily non-smokers and light-smokers. But the majority in the disease smoked more than ten cigarettes per day.

Approximately 50% of the participants in the disease group were from rural areas and 50% from urban areas. Most participants were from urban regions (Figure 7).

**Figure 7:**
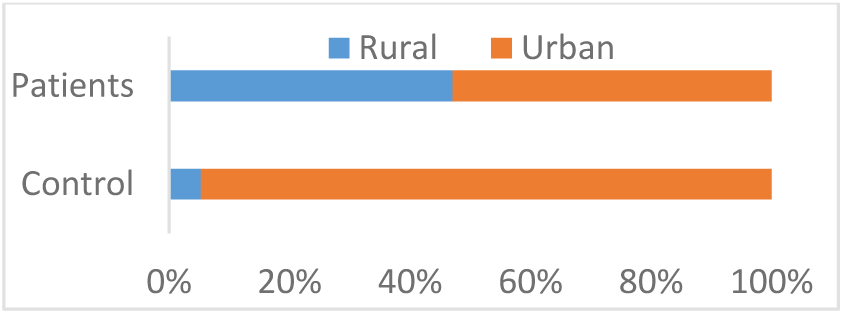
Place of living.

### Plasma IL-6 levels

In the control and disease groups, the mean plasma IL-6 levels were 0.97 pg/mL and 7.42 pg/mL, respectively (Figure 8). The difference was statistically very highly significant.

**Figure 8:**
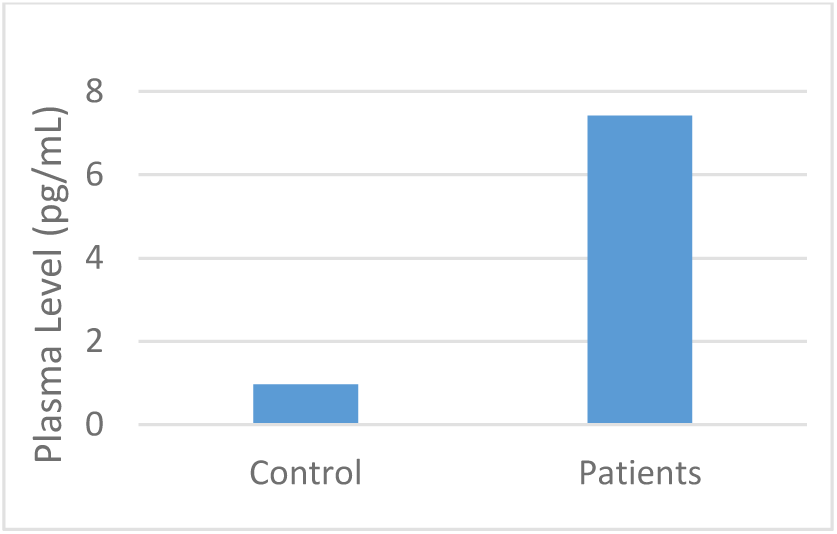
Plasma IL-6 levels in two groups.

There was no apparent difference in IL-6 levels in the control arm in males and females. However, the level was significantly higher in females than males in the diseased arm (Figure 9).

**Figure 9:**
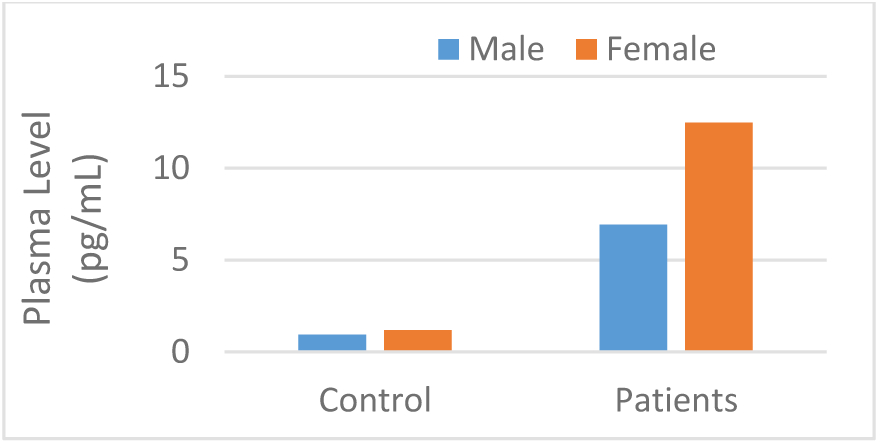
IL-6 levels vs. gender.

IL-6 levels were slightly higher in the age group of 41-60 years in lung cancer patients (Figure 10).

**Figure 10:**
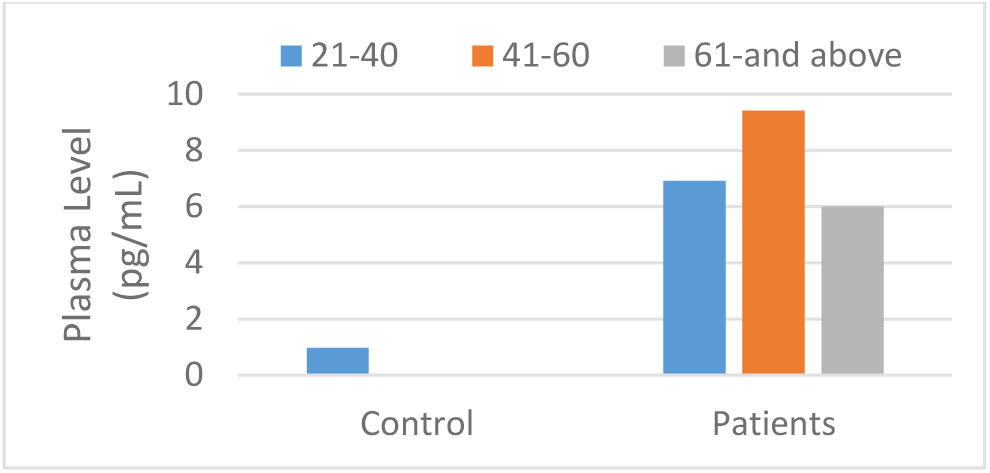
IL-6 vs. age groups.

Most lung cancer patients were diagnosed with either adenocarcinoma or squamous cell carcinoma in our study. The plasma IL-6 levels were slightly higher in adenocarcinoma patients (Figure 11).

**Figure 11:**
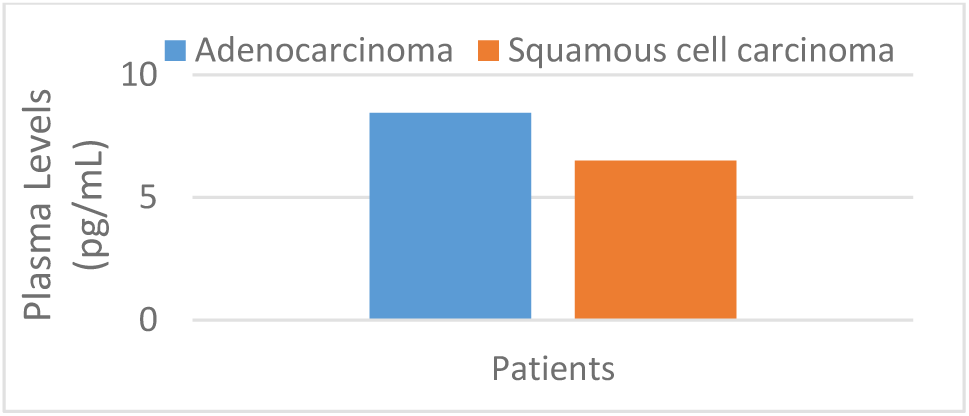
IL-6 levels vs. lung cancer subtypes.

Some patients received no anticancer therapy during recruitment to this study, some received only chemotherapy, and the remaining received both chemotherapy and radiotherapy. We did not find any difference in IL-6 levels depending on the treatment status (Figure 12).

**Figure 12:**
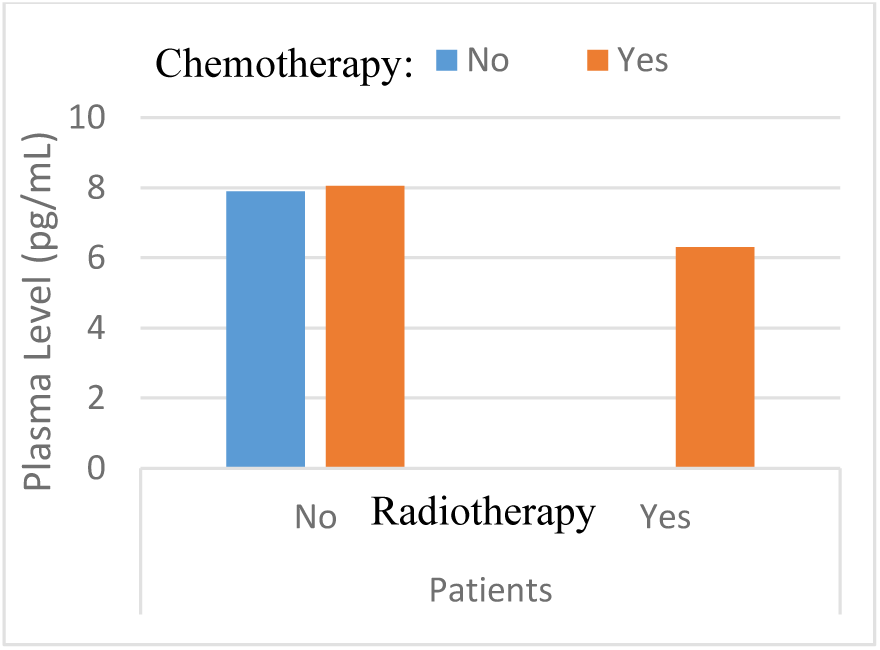
IL-6 levels in different treatment groups.

### Plasma IL-17 levels

In control and disease groups, the mean plasma IL-17 levels were 12.4 pg/mL and 9.4 pg/mL, respectively. Interestingly, IL-17 levels were strikingly higher in males than females in both groups (Figure 13).

**Figure 13:**
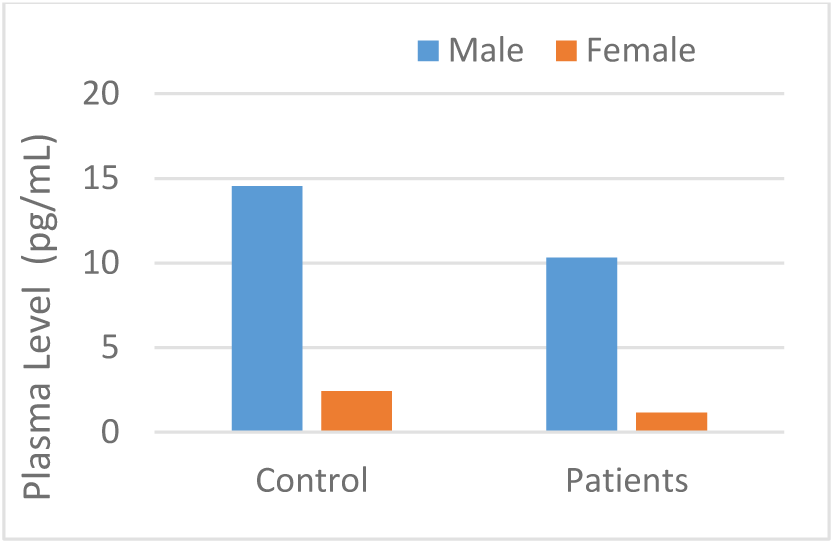
IL-17 levels vs. gender.

Our data indicate that plasma IL-17 concentrations were significantly higher in younger patients than patients 41 years and over (Figure 14).

**Figure 14:**
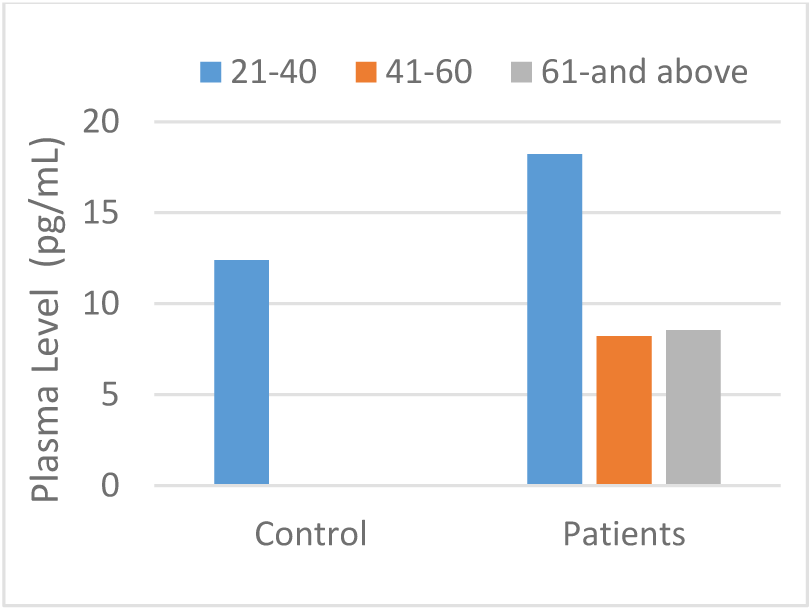
IL-17 levels vs. age groups.

**Figure 15:**
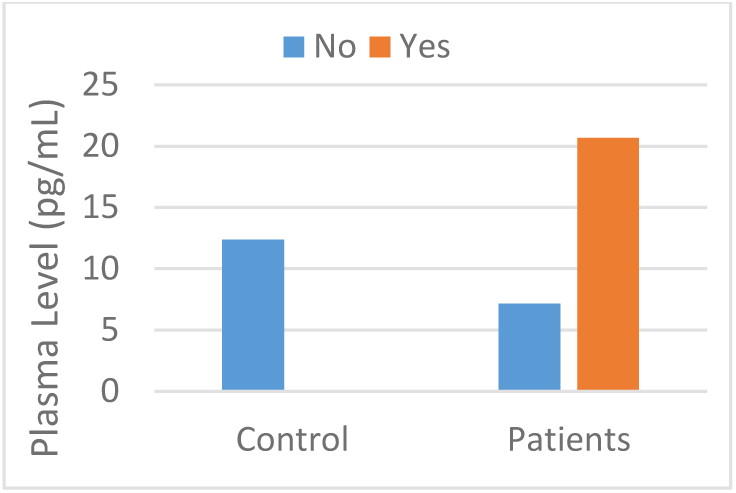
IL-17 levels vs. diabetes.

Interestingly, plasma IL-17 concentrations were significantly elevated in lung cancer patients with type 2 diabetes as a comorbidity (Figure 17).

## Conclusion

Our study confirmed previous works showing higher IL-6 plasma levels in lung cancer patients. Overall, we did not find any difference in IL-17 plasma levels between control and lung cancer patients. Interestingly, among the lung cancer patients, IL-17 level was considerably higher when diabetes was present as a comorbidity. Further analysis with larger samples will help establish the roles of interleukins in lung cancer diagnosis and prognosis.

## Data Availability

All data produced in the present study are available upon reasonable request to the authors

